# Bias-corrected serum creatinine from UK Biobank electronic medical records generates an important data resource for kidney function trajectories

**DOI:** 10.1101/2023.12.13.23299901

**Authors:** Mathias Gorski, Simon Wiegrebe, Ralph Burkhardt, Merle Behr, Helmut Küchenhoff, Klaus J. Stark, Carsten A. Böger, Iris M. Heid

## Abstract

**BACKGROUND:** Loss of kidney function is a substantial personal and public health burden. Kidney function is typically assessed as estimated glomerular filtration rate (eGFR) based on serum creatinine. Emerging electronic Medical Records (eMR) in UK Biobank present a promising resource to augment the data on longitudinal eGFR based on study center visits (SC; n=15,000). However, it is unclear whether eMR-based creatinine values can be used for research on eGFR trajectories.

**METHODS:** We derived eMR-based serum creatinine values (various assays/labs, Jaffe or enzymatic) from UK Biobank “GP-clinical”. We compared these with SC-based creatinine in individuals with both measurements available in the same calendar year (n=70,231; 2007-2012).

**RESULTS:** We found a multiplicative bias for eMR-based creatinine that was large, factor 0.84, for 2007, and decreased over time (0.97 for 2013). Deriving eGFR based on SC- and bias-corrected eMR-creatinine (CKD-Epi 2021) yielded 454,907 individuals with ≥1eGFR assessment (2,102,174 assessments). This included 206,063 individuals with ≥2 assessments (median=6.00 assessments) for a time between 1^st^ and last assessment of up to 60.2 years (median time=8.7 years). We enriched the dataset with eMR-recorded kidney-relevant events from “GP-clinical” (Acute Kidney Injury, End stage Kidney Disease, Nephrectomy, Dialysis, Kidney Transplant, Pregnancy, and Diabetes). We illustrated the suitability of this data: e.g. we found an annual eGFR decline of 1.04 mL/min/1.73m²/year (95%-CI=1.03-1.05), in line with literature and a four-fold steeper decline following Acute Kidney Injury.

**CONCLUSIONS:** In summary, our bias-correction of eMR-based creatinine values enabled a 4-fold increase in the number eGFR assessments in UK Biobank suitable for kidney function research.

## BACKGROUND

Accelerated kidney function decline can lead to renal failure, which necessitates dialysis or kidney transplantation. While age-related decline in kidney function is expected as part of the natural aging process (*1*), the rate of decline is highly variable in the population (*2*). The underlying reasons for this heterogeneity are not well understood. To investigate these reasons, large datasets on kidney function over time in the general population and recorded kidney-relevant clinical events are needed. However, such longitudinal datasets are sparse.

The UK Biobank (*3*) provides an opportunity to advance our understanding of kidney function decline in a general adult population. With its vast and diverse cross-sectional dataset from the study center (SC) assessments, encompassing over 500,000 participants, it provides an unprecedented resource for studying cross-sectional kidney function. However, longitudinal data on biomarkers over time, including kidney function biomarkers, is currently limited to ∼15,000 individuals assessed at SC 4 years after baseline. The emergence of electronic Medical Record (eMR) data from general practitioners (GP) for UK Biobank participants offers the opportunity to augment the existing SC data with longitudinal information. This data was released in September 2019 including multiple serum creatinine measurements over time and records of severe kidney events. Such eMR-based measurements are potential subject to various sources of bias (*4*). This can be technical measurement bias from heterogeneous laboratories and assays. This can also be selection bias, when individuals with eMR-based data including creatinine measurements are more prone to disease than individuals without eMR data. The usability of this eMR-based data to assess kidney function and its potential for meaningful integration with the SC data is yet to be fully explored. We estimated the glomerular filtration rate (eGFR) based on serum creatinine levels (*5*).

Our main objective was thus to augment the existing UK Biobank data on SC-based eGFR by eMR-based data from GPs (“gp-clinical”, application number 20272) to provide a longitudinal data resource for studying kidney function decline. Specifically, we (i) extracted and quality-controlled the GP-eMR data on serum creatinine, (ii) compared it with SC-based creatinine values; (iii) derived eGFR for the combined creatinine values and included recorded kidney-relevant events, and (iv) assessed the usability of this combined data for studying kidney function decline.

## MATERIAL AND METHODS

### UK Biobank data from the Study Center assessment and GP-clinical

UK Biobank data and SC assessment at baseline and, for a smaller subset of participants, at a 4-year follow-up was described previously (*3*). Briefly, UK Biobank is a prospective cohort study that included approximately 500,000 individuals aged 40-69 years at baseline recruited at 22 study centers in the United Kingdom. The SC assessment involved collecting participants’ blood samples and storing aliquots frozen at −80°C for further analysis (*6*).

Creatinine was measured in serum for all individuals from SC blood drawn at baseline and follow-up in a central laboratory according to standardized protocols (Enzymatic Beckman Coulter AU5800). We obtained these SC-based serum creatinine values (crea_SC_), the date of the SC visit baseline and follow-up (date-of-exam), age at that date (age-at-exam), and sex of participants (data fields 30700, 31, 34, 52 and 53).

Based on the UK Biobank “GP-clinical” table, we obtained raw creatinine values (crea_eMR_) via read codes (*7*) (**Supplementary Table 1**, details **Supplementary Note 1**).

### Ascertaining the comparability of crea_SC_ and crea_eMR_

Next, we merged crea_eMR_ to crea_SC_ for each person by date of blood draw (“date-of-exam”). Crea_eMR_ were measured by different laboratories and laboratories were starting to implement standards around the year 2009. Therefore, we investigated the possibility of a systematic technical bias in crea_eMR_ values that decreased the closer the measurement date approached 2009 or some years after (assuming a heterogeneous onset of use of standardized products). For this, we compared crea_SC_ and crea_eMR_ distributions, values, and via Bland-Altman-plot (*8*) focusing on individuals with both from the same calendar year (using crea_eMR_ closest in time to SC blood draw).

Considering crea_SC_ as gold standard (centralized laboratory; highly standardized protocol; using an Enzymatic Beckman Coulter AU5800), we evaluated whether crea_eMR_ exhibited a systematic bias: we assumed normally distributed crea_SC_ and crea_eMR_ on the log-scale, *X* := ln (*crea_sc_*)∼ *N* (*μ, σ*^2^) and *X*^*^:= ln (*crea_eMR_*)∼ *N* (*μ*\*, *σ*^*2^), respectively. We also assumed an additive error consisting of a random component, 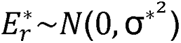, and a systematic bias, *s*,

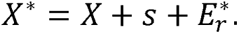

This implies that the expected value of 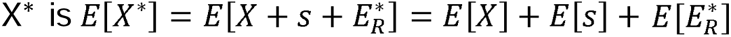, so that 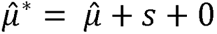, yielding 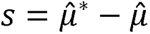. On the original scale, we can derive the geometric means of crea_eMR_ and crea_SC_, exp (*μ*\*) and exp (*μ*), respectively. Then the bias is multiplicative and given as the ratio of these geometric means

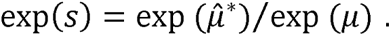

We assumed a differential bias by calendar years (larger in earlier years). Thus, we quantified the multiplicative bias (on the original scale) per calendar year, exp (*s^year^*) = exp (*μ*^*year^) /exp (*μ^year^*), among participants with crea_eMR_ and crea_SC_ values from the respective calendar year.

### Bias-corrected crea_eMR_

The above stated error model and bias quantification can also be used to derive bias-corrected crea_eMR_,

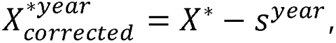

where *year* is the calendar year of the crea_eMR_ measurement. The expected value of the corrected crea_eMR_ is then 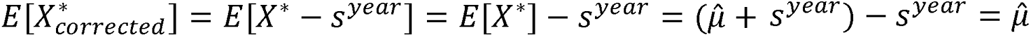, thus yielding 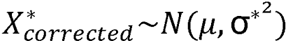, with a purely random error compared to crea_SC_, 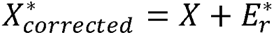.

For calendar years without individuals that had both crea_eMR_ and crea_SC_ available to estimate the bias correction factor, we used 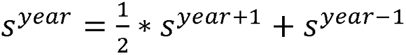, or *s^year^* from the first or last year, respectively. We utilized bias-corrected crea_eMR_ for further analyses, and merged them to crea_SC_ by person and date-of-exam.

### Generating the eGFR data based on crea_SC_- and crea_eMR_ values

We utilized the combined data of crea_SC_ and bias-corrected crea_eMR_ to derive eGFR (*5*). For this, we used age-at-exam (i.e. difference between date-at-exam and date-of-birth) and sex (reported at baseline SC visit). We defined Chronic Kidney Disease (CKD) as eGFR <60 mL/min/1.73m²).

### Integrating GP-eMR variables on kidney-relevant events

Next, we extracted GP-eMR data on kidney-relevant events: (i) onset of severe kidney events (End-Stage Kidney Disease, ESKD, Nephrectomy; Acute Kidney Injury, AKI), (ii) onset of renal replacement therapy (dialysis, kidney transplantation), (iii) onset of other conditions potentially relevant to kidney function (diabetes, pregnancy; **Supplementary Table 1**, **Supplementary Note 1**).

### Statistical Analyses

For our final dataset including kidney-relevant events and eGFR based on crea_SC_ or bias-corrected crea_eMR_, we derived descriptive statistics of the included UK Biobank study participants using SC-baseline information regarding lifestyle (smoking, BMI), diabetes (HbA1c>6.5%, self-report or medication) and CKD status (SC-based eGFR). Individuals with multiple eMR-based data points were reported to be potentially more affected by diseases (*4*). We thus examined whether individuals in GP clinical (GP clinical members) versus not in GP clinical, or, among GP clinical members with ≥9 versus 2-8 eMR-based eGFR assessments differed regarding lifestyle, diabetes or CKD status.

To investigate whether the derived data was suitable for research on eGFR decline, we estimated annual eGFR decline (and 95%-CI) as difference between last and 1^st^ eGFR assessment divided by the number of years in-between. For this, we restricted to individuals with ≥2 eGFR assessments at least one year apart, censoring eGFR values after onset of kidney-relevant events (for pregnancy, excluding values ±6 months before and after). We also estimated annual eGFR decline (and 95%-CI) around an incident AKI event using eGFR assessments ≥6 months (as close as possible) before and after AKI.

## RESULTS

### Serum creatinine measurements from the SC visits and GP-eMR

When extracting crea_SC_ from the baseline SC visit (year of exam 2006-2010), we yielded measurements for 425,147 individuals. For 16,446 individuals, crea_SC_ was available from the follow-up SC visit (year of exam 2012-2013). Together, this resulted in 15,314 individuals with 2 creatinine values over time for follow-up time of up to 6.11 years in-between (median time= 4.42 years) and 410,965 individuals with exactly one creatinine value.

When extracting crea_eMR_ from “GP-clinical” (*7*), we yielded 1,701,710 raw creatinine values for 199,482 individuals. After quality control, this resulted in 1,660,581 crea_eMR_ values for 199,968 individuals (**Supplementary Figure 1**). This included 23,188 individuals with exactly 2 creatinine measurements and 151,728 with ≥3 measurements (median number of measurements per person=7.00, max=288). The year of exam (i.e. year of measurement) was as early as 1950 up to 2017 resulting in a time between 1^st^ and last measurement of up to 60.15 years (median time=8.12 years).

When merging the crea_eMR_ values to the crea_SC_ values by date-of-exam (i.e. date of measurement for eMR; date of SC-visit for SC), we yielded 2,102,174 creatinine values for 454,907 individuals. Thus, the crea_eMR_ values substantially extended the longitudinal information on eGFR for UK Biobank participants including eGFR trajectories for up to 60.10 years and up to 289 measurements per person (**Figure 1A&B**; among the 206,063 individuals with ≥2 measurements: median time between 1^st^ and last assessment=8.71 years; median number of measurements per person=6.00).

**Figure 1:**
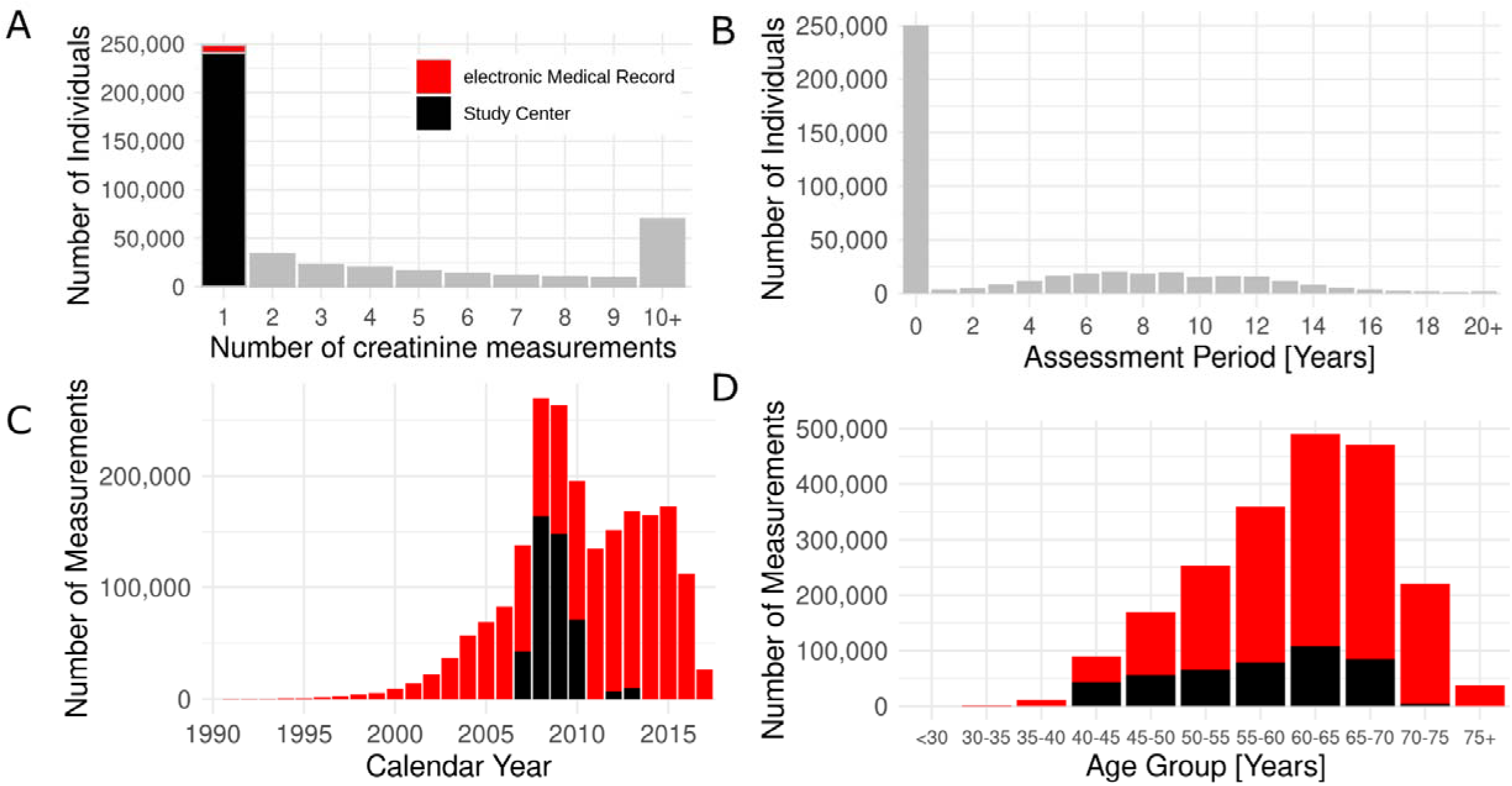
Descriptives of creatinine values from Study Center (SC) and electronic Medical Records (eMR). When combining SC- and eMR-based creatinine values after quality-control, we obtained 2,102,174 creatinine values from 454,907 individuals. We show (**A**) the distribution of individuals by their number of available creatinine measurements over time (n=248,844 with exactly one measurement indicated as SC- or eMR-derived in black or red, respectively; n=206,063 with ≥2 measurements ove time in gray), (**B**) the distribution of individuals by the time between 1^st^ and last measurement, (**C**) the number of measurements assessed in each calendar year, and (**D**) the number of measurements available by age groups (age-at-exam, “exam” referring to the GP-record for eMR or to the examination at the SC visit). For **C**&**D**, the red colour represents eMR and black colour represents SC data.

### Bias corrected crea_eMR_ values

We evaluated whether crea_eMR_ values were comparable to crea_SC_ data. We hypothesized that crea_eMR_ measurements conducted in years substantially earlier than 2009 were subject to a measurement bias. We further hypothesized that this bias became smaller the closer the measurement year was to 2009 and beyond, where laboratories started implementing standards for creatinine measurements (*9*). To investigate this, we focused on the 70,231 individuals with crea_SC_ and crea_eMR_ values in the same calendar year (year of exam 2007-2010 and 2011&2012). We observed higher median crea_eMR_ values compared to crea_SC_ values in earlier years, which gradually converged towards nearly identical median values in 2013, **Figure 2A, Supplementary Figure 2**).

**Figure 2:**
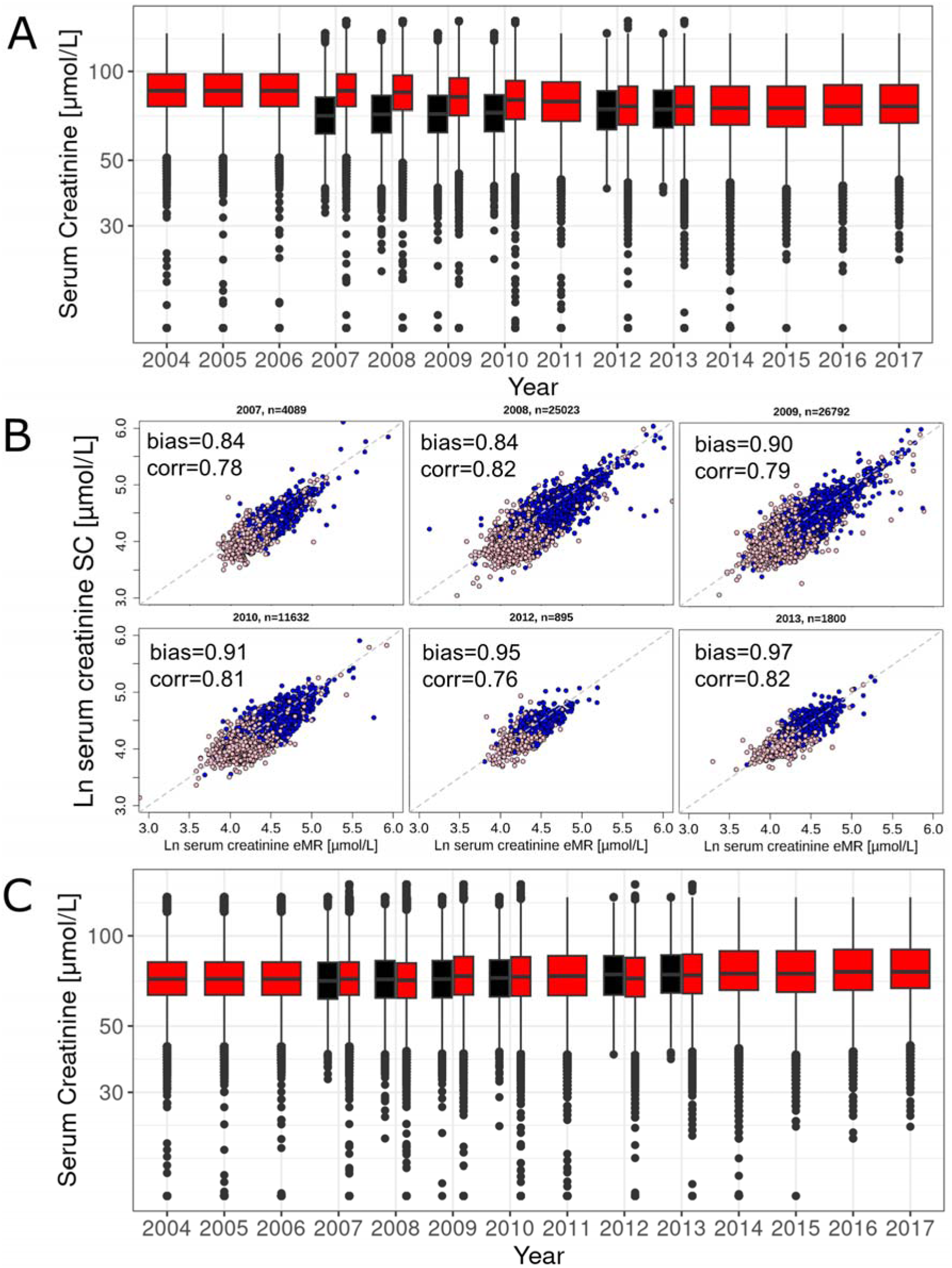
Bias correction of serum creatinine values from electronic Medical Records (eMR) data. We contrasted serum creatinine values from study center (SC) versus eMR by calendar year of exam (i.e. year of blood draw & measurement for eMR, year of blood draw for SC with centralized measurement in 2007-2010 and 2012-2013). (**A**) We show the distributions of creatinine from eMR (red; quality-controlled, not bias-corrected) and SC (black). (**B**) We show creatinine values from SC-versus eMR (quality-controlled, not bias-corrected) among 70,231 individuals with both measurements from the same calendar year (using the eMR-creatinine value closest in time to the SC-value). Also shown is the estimated bias (i.e. ratio of geometric means of SC-values versus eMR-values) and Spearman correlation coefficient. Grey lines indicate the identity. Sex is color-coded (blue: men, pink: women). (**C**) Shown are the distributions of bias-corrected creatinine from eMR (green; bias-corrected) and SC (black). The year-specific bias estimate was used as correction factor; for other years, we used proxies (2011: 0.93 derived as exp(average of ln(bias) for 2010 and 2012; years before 2007: 0.84; years after 2013: 1.0 (no correction).

We quantified the bias of crea_eMR_ based on an additive error model on the log-scale as the difference between mean log (crea_eMR_) and mean log (crea_SC_), *s*. This can also be interpreted as a multiplicative error on the original scale, where the bias is quantified as factor, *exp(s)*, derived as ratio of the geometric means of crea_eMR_ and crea_SC_ (**Methods**). We found a bias of factor 0.84 for the year 2007, which converged to near unity until year 2013 (0.84, 0.84, 0.90, 0.91, 0.95 and 0.97), for the years 2007-2010 and 2012-2013, respectively, **Figure 2B**). We obtained nearly the same bias estimates when winsorizing extreme values (beyond mean ± standard deviation on log-scale: exp(s) =0.84, 0.84, 0.90, 0.91, 0.95 and 0.98), emphasizing the lack of influence of extreme values.

In order to obtain bias-corrected crea_eMR_ values for all individuals, we used the derived year-specific correction factors exp (*s^year^*) to correct crea_eMR_ measurements from the respective year (i.e. 2007-2010, 2012 and 2013). For years where the correction factor could not be estimated directly, we used proxy correction factors: (i) the average of 2010 and 2012 on log-scale for 2011 (0.93); (ii) the 2007 factor for measurements before 2007 (0.84), (iii) and 1.0 (no correction) for measurements after 2013. When comparing the year-specific distributions of corrected crea_eMR_ with crea_SC_ distributions, we now found similar distributions and median values (**Figure 2C**).

### Description of the resulting UK Biobank dataset on eGFR including eGFR trajectories

Next, we merged crea_SC_ and bias-corrected crea_eMR_ values by date-of-exam and derived the eGFR (CKD-EPI 2021 (*5*)). When comparing the eGFR_eMR_ with the eGFR_SC_ values, we observed the bias before the correction and no bias after the correction (**Supplementary Figure 3&4**). We found that eGFR_eMR_ was comparable with eGFR_SC_ when regarded as 5-year age groups (**Figure 3A**).

**Figure 3:**
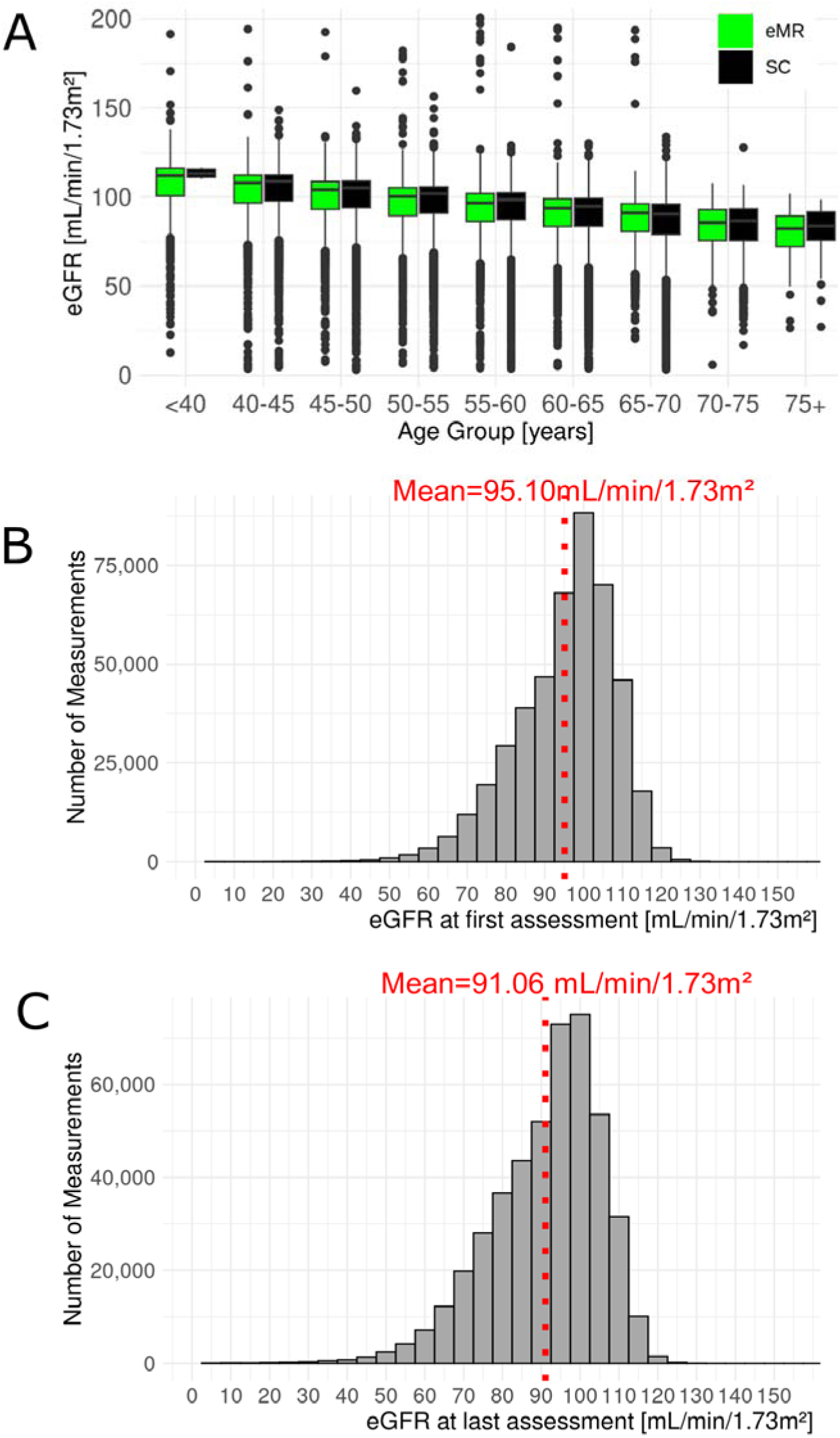
Descriptive of combined data from eMR- and SC-based eGFR. The dataset for eGFR from SC and eMR combined comprised 454,907 individuals with ≥1 eGFR assessment and overall 2,102,174 assessments of eGFR. (**A**) Shown are age-group-specific eGFR distributions (*5*) based on data from Study Center (SC, black) and electronic Medical Records (eMR; bias-corrected, green). This was limited to the 1^st^ eGFR assessment from SC or eMR, respectively. We also show eGFR distributions at **(B)** first and **(C)** last assessment (omitting 48 extreme values ≥ 150mL/min/1.73m²).

The final data comprised 454,907 individuals with ≥1 eGFR assessment based on crea_SC_ and/or crea_eMR_ (54.18% women) and with overall 2,102,174 eGFR assessments (**Table 1**). Average age was 55.9 years at the 1^st^ and 59.8 years at the last eGFR assessment. Mean eGFR was 95.10 and 91.07 mL/min/1.73m² at 1^st^ and last assessment, respectively (**Figure 3B&C**). The data consisted of (i) 248,844 individuals with exactly 1 eGFR assessment, mostly from SC baseline visit (98.26% from SC baseline, 0.32% from SC follow-up, 2.21% from eMR), (ii) 33,851 individuals with exactly 2 assessments (60.0% of these with one assessment from SC and one from eMR), and (iii) 172,212 with ≥3 assessments (i.e. ≥k-2 assessments from eMR, k being the number of assessments per person).

**Table 1:** Participant descriptive of the UKBB data on eGFR including eGFR trajectories. We show participant characteristics using the information from the study center (SC) visit at baseline as well as temporal aspects of the combined SC- and (bias-corrected) eMR-based data on eGFR (*5*). These descriptive statistics are shown for all individuals and with more than 2 and more than 10 eGFR assessments that can be used for analyses of eGFR trajectories (distinct subgroups with eGFR in individuals not in eMR data, with 1-8 and with ≥9 eMR-based data on eGFR in **Supplementary Table 2**). In the table header, ‘n’ denotes the number of individuals and ‘m’ denotes the number of eGFR assessments. Presented are mean and standard deviations, if not stated otherwise.

| | All Data<br>(n=454,907,<br>m=2,102,174) | $\geq 2$ eGFR<br>(n=206,063,<br>m=1,853,330) | $\geq 10$ eGFR<br>(n=69,506,<br>m=1,234,508) |
| --- | --- | --- | --- |
| **Characteristics from SC-baseline\*** |  |  |  |
| Age - years | 57.3 ± 8.1 | 57.9 ± 7.9 | 60.7 ± 6.9 |
| Sex – female (%) | 246,480 (54.2) | 111,528 (54.1) | 34,389 (49.5) |
| Ancestry - European (%) | 414,097 (91.0) | 180,359 (87.5) | 59,909 (86.2) |
| Smoking status **– current or ever (%) | 204,051 (44.9) | 93,244 (45.3) | 34,771 (50.1) |
| BMI **– kg/m<sup>2</sup> | 27.4 ± 4.8 | 27.6 ± 4.9 | 29.1 ± 5.3 |
| Diabetes – yes (%) | 19,479 (4.5) | 9,579 (4.9) | 8,219 (12.6) |
| eGFR - mL/min/1.73m<sup>2</sup> | 94.1 ± 13.2 | 93.6 ± 13.2 | 90.5 ± 14.5 |
| Chronic Kidney Disease**** – yes (%) | 7,258 (1.6) | 3,437 (1.8) | 2,284 (3.6) |
| **Temporal aspects** |  |  |  |
| Age-first-exam*** - years | 55.9 ± 8.2 | 54.7 ± 8.0 | 55.7 ± 7.3 |
| Age-last-exam*** - years | 59.8 ± 8.9 | 63.44 ± 8.5 | 67.6 ± 6.9 |
| time between 1<sup>st</sup> and last eGFR assessment - median (max) - years | 0.0 (60.2) | 8.5 (60.2) | 11.7 (60.2) |
| Calendar year of exam*** - min to max | 1950 to 2017 | 1950 - 2017 | 1950 – 2017 |
| **eGFR** |  |  |  |
| #eGFR assessments per person – median [IQR] | 1.0 [1.0 -5.0] | 6.0 [3.0-12.0] | 15.0 [12.0-20.0] |
| eGFR-first-exam - mL/min/1.73m<sup>2</sup> | 95.1 ± 12.9 | 95.9 ± 12.6 | 94.2 ± 13.3 |
| eGFR-last-exam - mL/min/1.73m<sup>2</sup> | 91.1 ± 14.4 | 86.9 ± 14.8 | 81.5 ± 16.5 |
| Chronic Kidney Disease**** – yes (%) | 23,035 (5.1) | 19,191 (9.3) | 14,212 (20.5) |

BMI=Body Mass Index, eGFR=estimated Glomerular Filtration Rate.
\* obtained from SC-baseline (data fields 30700, 31, 34, 52, 53, 20116, 21001, 30750, 20003, 2443 and 21001).
\*\* Among the individuals where this variable is available (n=454,361 for smoking status, n=452,721 for BMI).
\*\*\* "Exam" is the examination in SC or the examination by the GP, assuming that the date of the eMR-record is the same date as the GP-exam.
\*\*\*\* Chronic Kidney Disease was defined as having at least one assessment of eGFR <60 mL/min/1.73m<sup>2</sup>

The characteristics of individuals with ≥2 eGFR assessments (age, %women, %smoking, BMI, eGFR, and %CKD from SC-baseline) were similar as in the overall data, but individuals with ≥10 eGFR assessments were older, with higher BMI, lower eGFR, higher %CKD (**Table 1**). This difference between individuals with many versus few assessments was also observed when restricting to individuals that were part of GP-clinical data (GP-clinical members), while there was no difference between GP-clinical members and individuals that were not in GP-clinical (n=199,396 versus 255,511, respectively; **Supplementary Table 2**). Thus, we found evidence for a selection towards older and less healthy individuals among those with many crea_EMR_ values over time compared to fewer crea_eMR_ values, in line with literature (*4, 10*). However, there was no selection observed for being a GP-clinical member versus all UK Biobank participants.

### Description of eGFR trajectories augmented with kidney-relevant clinical events in the resulting UK Biobank dataset

To determine eGFR trajectories in the UK Biobank dataset, we analyzed 206,063 individuals with ≥2eGFR assessments over time, finally encompassing 1,853,330 eGFR assessments as UK Biobank dataset on eGFR trajectories (**Table 1**). Most of these individuals (n=195,885, 95.1%) had ≥1 eGFR assessed from crea_eMR_, thus were members of GP-clinical. Median time between 1^st^ and last assessment was 8.5 years (IQR: 5.8-11.5; maximum 60.2 years), median number of eGFR assessments over time per person was 6.0 (IQR: 3.0-12.0; up to 289 assessments).

A key aspect when analysing eGFR decline over time is the censoring (i.e. set to missing) of eGFR values at or after severe kidney events (ESKD, AKI, nephrectomy), after onset of renal replacement therapy (dialysis, kidney transplantation), and during or shortly after pregnancy. We identified read codes for records of AKI, ESKD, Nephrectomy, Dialysis, Kidney Transplantation and Pregnancy, curated based on experts’ knowledge, and we obtained read codes for Diabetes (*11*) (**Supplementary Table 1**). We merged the corresponding information from UK Biobank “GP-clinical” to the eGFR trajectories by date-of-event compared to the date-of-exam for the eGFR assessment (**Figure 4, Supplementary Table 3**). Among the 199,396 individuals with ≥2 eGFR value over time that were GP-clinical members (i.e. ≥1 eMR-based eGFR value), we recorded >1000 kidney-relevant events (**Supplementary Table 3**). For example, 94 individuals had AKI before the 1^st^ eGFR assessment and 563 had an AKI event after the 1^st^ eGFR assessment. We compared the 563 individuals with a recorded AKI event to the 171,565 individuals with no recorded AKI event. Individuals with AKI also had more eGFR assessments over time (median=4.0, IQR: 1.0-14.0) compared to those without AKI (median=1.0, IQR: 1.0-5.0). Overall, there were more individuals with a severe kidney event or onset of renal replacement therapy among the individuals with ≥10 eGFR assessments than among the GP-clinical members (i.e. ≥1 eMR-eGFR assessment)

**Figure 4:**
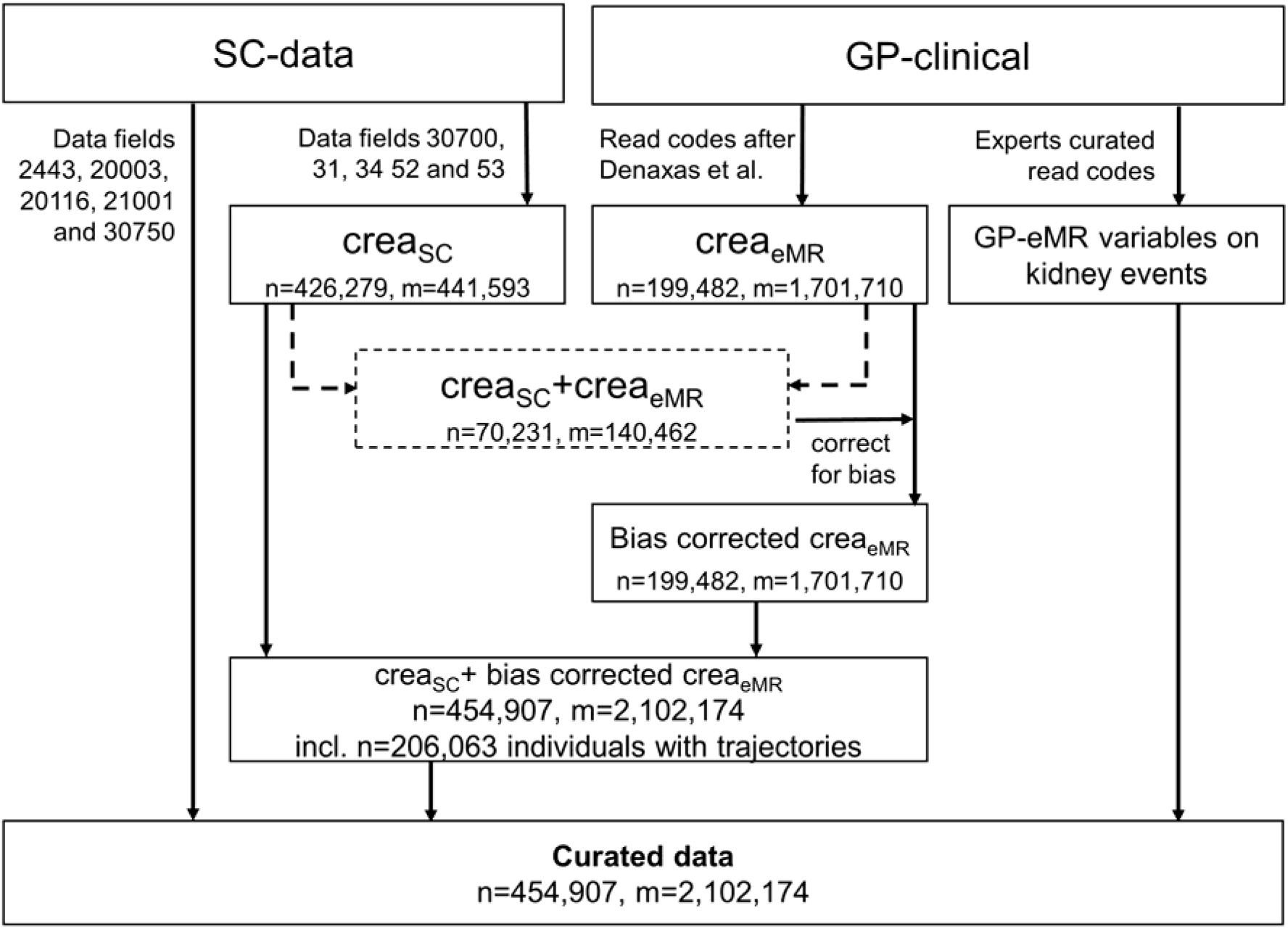
Workflow from raw data to the curated data. This schematic illustrates the analytical flow of the data generation. We used Study Center (SC) data on creatinine at baseline and follow-up (crea_SC_, data field 30700) and other SC-based data (date-of-exam, age-at-exam, sex, BMI, smoking status, HbA1c, medication and diabetes diagnosis; data fields 53, 34, 52, 31, 21001, 20116, 30750, 20003, 2443). We used eMR-data from “GP-clinical” deriving reads codes for creatinine (crea_eMR_) (*7*), Diabetes (*11*), and read codes for kidney-function related events and data-of-exam (Acute Kidney Injury, End stage Kidney Disease, Nephrectomy, Dialysis, Kidney Transplant and Pregnancy). We derived the calendar-specific bias correction factor for crea_eMR_ in individuals with both crea_eMR_ and crea_SC_ available in the same calendar year (dashed lines) and used proxy correction factors for other calendar years. We combined crea_SC_ and bias-corrected crea_eMR_ with other SC-based data and eMR-based events. ‘n’ denotes the number of individuals and ‘m’ the number creatinine measures.

### Some aspects of UK Biobank eGFR trajectories exemplifying its utility

One aspect to provide a proof-of-concept that the data is usable to study eGFR decline is the annual decline observed in the data. When restricting to the 206,063 individuals with at least 2 eGFR assessments, a follow-up time of at least 1 year, and the censoring of eGFR values as stated above, we estimated annual decline (difference between last and 1^st^ eGFR assessment divided by years in-between) of 1.04 mL/min/1.73m² per year (95%-CI=1.03-1.05). This is in line with literature (*1*) and documented a plausible eGFR decline in the data despite heterogeneous source of creatinine measurements. Of note, when using the crea_eMR_ values without bias-correction, the mean annual eGFR decline was 0.11 mL/min/1.73m² per year (95%-CI=0.10-0.12). This underscored the importance of the bias-correction when studying eMR-based eGFR from historic creatinine measurements in UK Biobank GP-clinical.

To provide a further proof-of-concept, we evaluated whether eGFR trajectories reflected properly the onset of severe kidney disease: e.g. for the 354 individuals with incident AKI and at least one eGFR assessment 6 months before and after, the mean annual eGFR decline was four times larger than the overall decline (3.93 mL/min/1.73m² per year, 95%-CI=2.82-5.05). An example of such a rapid eGFR decline around AKI is illustrated in **Figure 5A**. Two further examples illustrate eGFR trajectories of individuals with ESKD followed by transplantation or dialysis (**Figure 5B&C**).

**Figure 5:**
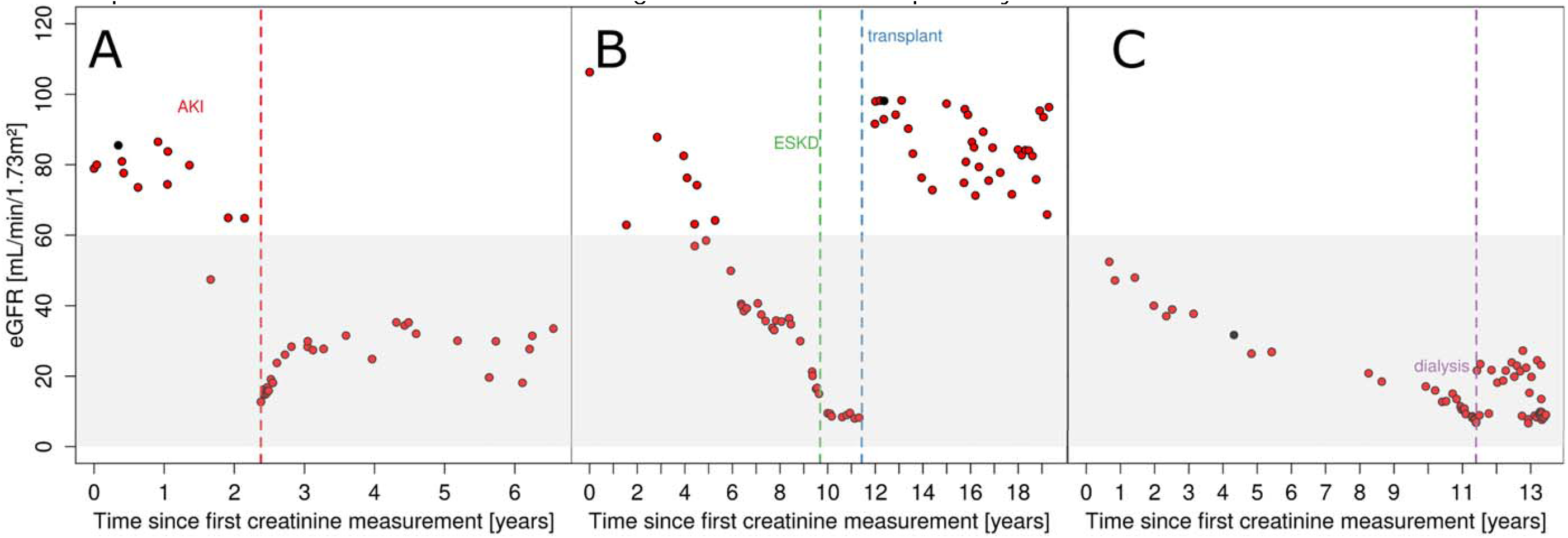
Showcases of eGFR trajectories for individuals with recorded kidney-relevant events. Shown are three examples of individuals: (**A**) a 65-year-old with an episode of Acute Kidney Injury (AKI) and subsequent partial eGFR recovery, (**B**) a 50-year-old with eGFR decline down to 10 mL/min/1.73m², a recording of ESKD, and eGFR recovery after kidney transplantation and (**C**) a 56-year-old with eGFR decline below 15 mL/min/1.73m² and under dialysis 11 years after the first eGFR assessment. The x-axis depicts the time since 1^st^ eGFR assessment in years, the y-axis depicts the eGFR value in mL/min/1.73m². Red and black dots indicated whether the eGFR was based on electronic Medical Records or from Study Center assessment, respectively. The shaded area indicates eGFR < 60 mL/min/1.73m² (i.e. defining Chronic Kidney Disease, CKD). Events of ESKD, AKI and transplant are visualized at vertical dotted lines in green, red and blue, respectively.

### Technical aspects of the UK Biobank data including eGFR-trajectories from eMR

In this dataset, we included all 450,000 individuals with ≥1 eGFR assessment, since some analytical approaches to analyze eGFR decline can integrate individuals with only one GFR assessment (e.g. linear mixed models). We generated the data as long format (individual-identifier and record-number as key variables): the record-number counts the number of entries per person that can be (i) the quality-controlled, bias-corrected crea_eMR_ or crea_SC_ value together with the record-type (crea) and record-date or (ii) the kidney-relevant event together with record-type (onset of AKI, ESKD, nephrectomy, dialysis, kidney transplantation, pregnancy). The integrated data utilizes SC data from September 2019 and GP-clinical downloaded in May 2023 (i.e. records until September 2017, depending on country Scotland, Wales or England).

The newly generated variables for this UK Biobank dataset (**Supplementary Table 4**) are available as a return dataset in the UK Biobank portal.

## CONCLUSIONS

With this work, we present quality-controlled curated UK Biobank data of 2,102,174 creatinine measurements and calculated eGFR in 454,907 individuals. For 206,063 of these individuals, eGFR trajectories are available with two or more eGFR assessments over time augmented with kidney-relevant events from GP-clinical. Thus, by including eMR information from GP-clinical, we extended the UK Biobank data on eGFR by >10-fold more individuals with longitudinal eGFR information and >2-fold longer time between 1^st^ and last eGFR assessment when compared to the dataset obtained in the SC, thus vastly extending the resource of kidney function in the UK Biobank.

A key aspect of eMR-based research on eGFR decline and thus a specific focus of our work was to determine the difference between crea_eMR_ and crea_SC_ measurements. The crea_eMR_ values stemmed from historic measurements in different laboratories and assays from routine outpatient care, and are thus prone to bias inherent to unstandardized creatinine assays. UK Biobank provided the opportunity to compare the crea_eMR_ with crea_SC_, which were all measured after SC visits on biobanked serum using a modern enzymatic creatinine assay from 2019 calibrated to National Institute Standardized Technology (NIST) reference material 967. We observed a bias in the eMR measurements of factor 0.84 for calendar year 2007 and 2008, which decreased to factor 0.90 for the year 2009, coinciding with implementing standards for creatinine measurements (*9*). The observed bias of 0.95 and 0.97 for the years 2012 and 2013 could reflect a slow and heterogeneous onset of laboratory standardization. Using the correction factor of 0.84 for years before 2007 and no bias-correction after 2013 might not be a perfect fit, but provides a best reflection of our data. Our proof-of-concept analyses estimating the annual eGFR decline without and with bias-corrected values showed 0.11 versus 1.04 mL/min/1.73m² per year annual decline, with the latter being in line with literature (*1*). Thus, we not only provided curated and bias-corrected eMR-based eGFR values for UK Biobank individuals that can be reasonably used for research on eGFR decline. We also provided an approach to identify, quantify, and account for bias in eMR-based biomarker, when gold standard measurements were available.

For studies with historic creatinine measurements but no gold standard measurements, this might not be fully generalizable. However, the limited standardization of serum creatinine measurements before 2009 and implemented standards thereafter were well acknowledged (*9*). It is conceivable that the here presented correction factors by calendar year might provide some reasonable proxy quantification of the bias involved also for other eMR-based studies. With all due concern regarding the problem of external data and limited generalizability to other settings, using the here presented correction factors might be better than no bias correction of historic serum creatinine measurements.

A limitation is the heterogeneity of crea_eMR_ values across laboratories and assays that probably involve larger measurement error than standardized measurements even after bias correction. Another limitation is the fact that the data is predominantly European ancestry, which is a general limitation of UK Biobank. Data from eMR are generally prone to bias with regard to enrichment of individuals with disease (*12*). While we found more AKI events among individuals with many versus few eMR-records on creatinine, we found little differences between UK Biobank participants that had eMR-records (i.e. were members of the GP-clinical) versus other participants. This might be due to the fact that UK GP-records are rather comprehensive.

In other recent work on datasets of eGFR trajectories, eGFR trajectories are utilized from 116,870 individuals with CKD from the Million Veterans Project and Vanderbilt University Medical Centers identifying novel genetic loci associated with longitudinal eGFR decline (*13*). The topic is timely as large-scale Biobank data that integrate eMR data on biomarker trajectories for research are currently emerging, but so far with little focus on serum creatinine and kidney function decline (e.g. all-of-us) (*11, 14*).

In summary, the quality-controlled and bias-corrected eMR-based information on eGFR enables analyses of eGFR trajectories in UK Biobank. We also provide an approach to identify, quantify, and correct for bias in historic creatinine measurements that is applicable to other biobank data. Further analyses of large data on eGFR trajectories together with rich information on genetic and non-genetic risk factors can help understand the variability in eGFR decline in the population.

## DATA AVAILABILITY

We will make the data available for download through the UK Biobank portal as a Return data set of the project number 20272. This data is available to researchers registered with the UK Biobank: please refer to instructions within the AMS portal to download these results.

## ACKNOWLEDGMENTS

This work was funded by the Deutsche Forschungsgemeinschaft (DFG, German Research Foundation), Project-ID 387509280, SFB 1350; Project-ID 509149993, TRR 374. This work was also supported by the German Federal Ministry of Education and Research (grant number BMBF 01ER1206, BMBF 01ER1507 to I.M.H.) and by the German Research Foundation (grant number HE 3690/7-1 to I.M.H). This work was conduct with UK Biobank application 20272. We thank the UK Biobank and all their participants.

## AUTHOR CONTRIBUTIONS

M.G. and I.H wrote the manuscript, designed the study and interpreted the results. R.B., C.B. and K.S. provided expertise on the quality control of creatinine measurements. I. H. and H.K. provided expertise for the bias correction, description and statistical analyses. S. W, H.K. and T.W. helped interpret the results. All authors critically reviewed the manuscript.

## ONLINE MATERIAL

The gp-clinical was downloaded in May 2023 and the accompanying material was downloaded from https://biobank.ctsu.ox.ac.uk/~bbdatan/biomarkers.pdf at the same date. Material accompanying the gp-clinical table on serum biochemistry were downloaded from biobank.ctsu.ox.ac.uk/ukb/ukb/docs/serum_biochemistry.pdf (Oct 12^th^ 2022)

## REFERENCES

1. Denic A, Glassock RJ, Rule AD. The Kidney in Normal Aging: A Comparison with Chronic Kidney Disease. Clin J Am Soc Nephrol 2022;17:137–9.

2. Levey AS, Becker C, Inker LA. Glomerular filtration rate and albuminuria for detection and staging of acute and chronic kidney disease in adults: a systematic review. JAMA 2015;313:837–46.

3. Sudlow C, Gallacher J, Allen N, Beral V, Burton P, Danesh J, et al. UK biobank: an open access resource for identifying the causes of a wide range of complex diseases of middle and old age. PLoS Med 2015;12:e1001779.

4. Agniel D, Kohane IS, Weber GM. Biases in electronic health record data due to processes within the healthcare system: retrospective observational study. BMJ 2018;361:k1479.

5. Inker LA, Eneanya ND, Coresh J, Tighiouart H, Wang D, Sang Y, et al. New Creatinine- and Cystatin C-Based Equations to Estimate GFR without Race. N Engl J Med 2021;385:1737–49.

6. Peakman TC, Elliott P. The UK Biobank sample handling and storage validation studies. Int J Epidemiol 2008;37 Suppl 1:i2–6.

7. Denaxas S, Shah AD, Mateen BA, Kuan V, Quint JK, Fitzpatrick N, et al. A semi-supervised approach for rapidly creating clinical biomarker phenotypes in the UK Biobank using different primary care EHR and clinical terminology systems. JAMIA Open 2020;3:545–56.

8. Bland JM, Altman DG. Statistical methods for assessing agreement between two methods of clinical measurement. Lancet 1986;1:307–10.

9. Coresh J, Turin TC, Matsushita K, Sang Y, Ballew SH, Appel LJ, et al. Decline in estimated glomerular filtration rate and subsequent risk of end-stage renal disease and mortality. JAMA 2014;311:2518–31.

10. Sauer CM, Chen L-C, Hyland SL, Girbes A, Elbers P, Celi LA. Leveraging electronic health records for data science: common pitfalls and how to avoid them. Lancet Digit Health 2022;4:e893–e898.

11. Ko S, German CA, Jensen A, Shen J, Wang A, Mehrotra DV, et al. GWAS of longitudinal trajectories at biobank scale. Am J Hum Genet 2022;109:433–45.

12. Young JC, Conover MM, Funk MJ. Measurement error and misclassification in electronic medical records: methods to mitigate bias. Curr Epidemiol Rep 2018;5:343–56.

13. Robinson-Cohen C, Triozzi JL, Rowan B, He J, Chen HC, Zheng NS, et al. Genome-Wide Association Study of CKD Progression. J Am Soc Nephrol 2023;34:1547–59.

14. Zhou W, Kanai M, Wu K-HH, Rasheed H, Tsuo K, Hirbo JB, et al. Global Biobank Meta-analysis Initiative: Powering genetic discovery across human disease. Cell Genom 2022;2:100192.

