## Supplementary Material for "Bias-corrected serum creatinine from UK Biobank electronic medical records generates an important data resource for kidney function trajectories"


| | |
|---|---|
| **Supplementary Note1** | Quality control of serum creatinine measurements from eMRs and extraction of kidney-relevant events from GP-eMR |
| **Supplementary Table 1** | Clinical Terms Version 2 (Read v2) and Clinical Terms Version 3 (CVT3) of events**.** |
| **Supplementary Table 2** | Participant descriptives of GP-clinical members versus non-members |
| **Supplementary Table 3** | Kidney function relevant events |
| **Supplementary Table 4** | Overview of variables for time-varying and constant records. |
| **Supplementary Figure 1** | Quality control of serum creatinine measurements from eMRs. |
| **Supplementary Figure 2** | Average between creatinine from Study Center (SC) and electronic Medical Records (eMR) |
| **Supplementary Figure 3** | Comparing eGFR values from Study Centrer ($eGFR_{SC}$) with eGFR-values from electronic Medical Records ($eGFR_{eMR}$) before correcting the creatinine values from electronic Medical Records. |
| **Supplementary Figure 4** | Comparing eGFR values from Study Centrer ($eGFR_{SC}$) with eGFR-values from electronic Medical Records ($eGFR_{eMR}$) after correcting the creatinine values from electronic Medical Records. |

**Supplementary References**

### Supplementary Note 1: Quality control of serum creatinine measurements from eMRs and extraction of kidney-relevant events from GP-eMR

Read codes, specifically Clinical Terms Version 2 (Read v2) and Clinical Terms Version 3 (CTV3), are standardized vocabularies to codify clinical terms. In the UK healthcare system, individuals typically consult a GP, who refers patients to specialized care or requests biomarker measurements when indicated. Serum creatinine measurements are then conducted in contracted laboratories. The specific assays and protocols used for these measurements are unknown and presumed to vary between laboratories and time points.

We quality-controlled the $crea_{eMR}$ values for technical artefacts: (i) we excluded zero values. (ii) For creatinine values obtained on the same day ("duplicates"), we used the mean of natural logarithm of the two measures when the two measurements were within a 10% tolerance, excluded a value that was 88.4 times higher than the other (i.e. same value coded both in md/dL and µmol/L), or excluded both otherwise. (iii) We excluded values below the level of detection (LOD, 0.88 µmol/L, 0.01mg/dL (*1*) or above the highest ever recorded value of 6524 µmol/L (73.8 mg/dL) (*2*). (iv) We set values below the level of quantification (LoQ, 4.42 µmol/L, 0.05 mg/dL (*1*) to the LoQ. This yielded our quality-controlled $crea_{eMR}$ values (in µmol/L) and the date of recorded measurement. We assumed that this date was equivalent or close to the data of blood draw (i.e. "date-of-exam", **Supplementary Figure 1**).

To extract GP-eMR data on kidney-relevant events, we searched in the Coding System Lookups and Mappings Dictionary (Version 3, May 2021) for kidney-relevant events terms and identified the corresponding Read v2 and CTV3 read codes (**Supplementary Table 1**). Then, we extracted the GP-clinical information for these read codes. We merged an individual's events data into the eGFR data according to the date-of-exam (i.e. date-of-exam for recorded event, date-of-exam for eGFR assessment).

For the GP-clinical released Sept 2019, the extraction date of eMR on behalf of the UK Biobank was between August 2016 and September 2017, depending on country (Scotland, Wales or England) and GP Computer System Supplier (EMIS, Vision or TPP). Thus, there is no serum creatinine value or kidney-relevant event after September 2017 in the presented data.

**Supplementary Table 1: Clinical Terms Version 2 (Read v2) and Clinical Terms Version 3 (CVT3) of events**.
For the here described events, we extracted read Codes from the dictionary "Coding system lookups and mappings - Version 3, May 2021." Read codes for Creatinine and Diabetes were extracted by the approach and the described read codes from Denaxas et al., while all others were ascertained through manual retrieval of the search terms of the Events (e.g. "acute kidney injury") for Read v2 or CTV3 read terms.

| Event | Read code V2 or CTV3 code |
|---|---|
| Creatinine | "44J3.","44J30","44J31","44J32","44J33","44J3z","44JD.","44JC.","44J32","44J3z","44J33","44J31","44J30","XE2q5","XaERc","XaERX" |
| AKI | “XE2QM”,"X30Is","XaZ6J","XaZPp","XaZPs","XaZPt","XaZPu","XaPwv","XaZSx","X30Ir","XaZUY","XaZUZ","XaZUa","XaZYv","XaZZ0","XaZZ2","XaZe5","Xaa8O","Xaa8P","Xaa8Q","Y31fg","Yavic","Y31fh","Y31fn","Y31fo","Y31fr","Y31gQ","Y31gN","Y31gO","Y31fu","YavK2","YavVI","YavVL","YavVM","YavVN","Yasal","YavX4","YavYI","YavYJ","YavYK","YavYL","YavbY","Yavbb","Yavbd","Yavgj","Yaw12","Yaw13","Yaw14","Y31fj","Y31fi","K04..","K040.","K041.","K042.","K043.","K0430","K0431","K0432","K0433","K0434","K044.","K045.","K046.","K0460","K0461","K047.","K048.","K049.","K04A.","K04B.","K04C.","K04D.","K04E.","K04y.","K04z." |
| ESKD | “K05..”,"K050.","K055.","X30J0","Y31gA","XaLHK","YaoLB" |
| Nephrectomy | “7B01.”,"7B01.","7B010","7B010","7B010","7B011","7B012","7B013","7B013","7B014","7B015","7B015","7B016","7B017","7B018","7B019","7B01y","7B01z","7B02.","7B02.","7B020","7B021","7B022","7B023","7B02y","7B02z","7B01.","7B01.","7B010","7B010","7B010","7B011","7B012","7B013","7B013","7B014","XE0Fv","Xa2h5","7B016","7B017","7B018","Xa2h5","7B01y","7B01z","7B02.","7B02.","7B020","7B021","7B022","7B023","7B02y","7B02z","Y30ez","Y30f0","Y30f3","Y30f3","Y30f4","Y30f6","Y30fA","Y30fM","Y30fN","Y30fB","Y30fD","Y30fE","Y30fF","Y30f7","Y30f8","Y30fE","Y30f2","Y30f1","Y30fH","Y30fI","Y30fL","Y30fO","Y30fQ","Y30fS","Y30fK","Y30fJ" |
| Dialysis | “.14V2”,"14V2.","Ya07N","XE0Jg","Xa8S7","Xa8S7","Ya0Wc","7L1A.","7L1A0","7L1A1","7L1A2","7L1A3","7L1A4","7L1A5","7L1A6","7L1A7","XE0Jf","Y75Sk","YMIKl","Ya15a","Y03dD","Y03dH","7L1A1","7L1A2","X01AL","XaM2A","X40c3","XaMMt","X01AV","Y42A4","YaV79","Y03dQ","YapAD","Y42A6","YapYh","Y03dh","Y03c7","8882.","Y74u7","TB11." |
| Kidney Transplant | “7B00.”,"7B000","7B001","7B001","7B002","7B002","7B002","7B002","7B002","7B003","7B004","7B005","7B006","7B00y","7B00z","XaM1o","XaM1p","XaMKM","X30D2","Y30ef","Y30ew","Y30eq","Y30er","Y30el","Y30ep","Y30el","Y30ep","Y30el","Yap9l","Yap9m","YapW3","Y30ev","Y30ek","Y30ej","TB001","Y74uZ","ZV420","Ya0O1" |
| Pregnancy | “615C.”,"Y79tL","Y79tM","62...","6217.","6218.","6219.","621A.","621B.","621C.","621D.","621Z.","Y7EF5","Y79Qh","Y79Qj","X74V6","Y79Qi","621..","Y7EF7","Y7EF6","6211.","Y7EFF","6212.","Y7EFE","6213.","Y7EFD","6214.","Y7EFC","6215.","Y7EFB","6216.","Y7EFA","Y7EEZ","Y7EEY","Y7EEN","Y7EEX","Y7EEU","X76Qo","Y7EEV","X40Ah","Y40j9","Y7EF9" |

| | |
|---|---|
| Diabetes | "1252.,"1434.","C10..,"C100.","C1000","C1000","C1000","C1001","C1001","C1001","C1001","C1001","C100z","C101.","C101.","C101.","C1010","C1010","C1011","C1011","C101y","C101z","C102.","C102.","C1020","C1021","C102z","C103.","C1030","C1030","C1031","C1031","C1031","C103y","C103z","C104.","C104.","C104.","C1040","C1041","C104y","C104z","C105.","C1050","C1051","C105y","C105z","C106.","C106.","C106.","C106.","C106.","C1060","C1061","C106y","C106z","C107.","C107.","C107.","C107.","C1070","C1071","C1072","C1073","C1074","C107y","C107z","C1080","C1080","C1080","C1081","C1081","C1081","C1082","C1082","C1082","C1083","C1083","C1083","C1085","C1085","C1085","C1086","C1086","C1086","C1087","C1087","C1087","C1087","C1088","C1088","C1088","C1089","C1089","C1089","C108y","C108z","C1090","C1090","C1090","C1091","C1091","C1091","C1092","C1092","C1092","C1093","C1093","C1093","C1094","C1094","C1094","C1095","C1095","C1095","C1096","C1096","C1096","C1096","C1097","C1097","C1097","C10A0","C10A1","C10A2","C10A3","C10A4","C10A5","C10A6","C10A7","C10B0","C10y.","C10y0","C10y1","C10yy","C10yz","C10z.","C10z0","C10z1","C10zy","C10zz","C11y0" |

AKI=Acute Kidney Injury, ESKD=End-stage Kidney Disease. We obtained the read codes for creatinine and Diabetes from Denaxas et al.

**Supplementary Table 2: Participant descriptives of GP-clinical members versus non-members**

We show participant characteristics using the information from the study center (SC) visit at baseline and temporal aspects of the combined SC- and (bias-corrected) eMR-based eGFR (*3*). This is shown for members and non-members of "GP-clinical" and, for GP-clinical members separately by individuals with 1-8 and ≥9 eMR-based eGFR assessments (i.e. comparable to individuals with 2-9 versus ≥10 eGFR assessments for eMR- and SC-based eGFR combined). 'n' denotes the number of individuals and 'm' the number of eGFR assessments. Presented are mean and standard deviations, if not stated otherwise.

| | **Non-members GP-clinical (n=255,511, m=265,689)** | **Members GP-clinical 1-8 eMR-eGFR (n=120,952, m=521,535)** | **Members GP-clinical ≥9 eMR-eGFR (n=78,444, m=1,314,950)** |
|---|---|---|---|
| **Characteristics from SC-baseline*** | | | |
| Age - years | 57.0 ± 8.2 | 55.8 ± 8.0 | 60.5 ± 7.0 |
| Sex – female (%) | 137,993 (54.0) | 69,280 (57.3) | 39,207 (50.0) |
| Ancestry - European (%) | 242,813 (95.0) | 103,725 (85.8) | 67,559 (86.1) |
| Smoking status **– current or ever (%) | 113,508 (44.5) | 51,605 (42.7) | 38,938 (49.7) |
| BMI **– kg/m² | 27.3 ± 4.7 | 26.8 ± 4.4 | 29.0 ± 5.2 |
| Diabetes – yes (%) | 10,249 (4.2) | 832 (0.7) | 8,398 (11.4) |
| eGFR - mL/min/1.73m² | 94.3 ± 13.1 | 95.9 ± 11.9 | 90.8 ± 14.4 |
| Chronic Kidney Disease**** – yes (%) | 4,176 (1.6) | 695 (0.7) | 2,387 (3.4) |
| **Temporal aspects** | | | |
| Age-first-exam*** - years | 56.8 ± 8.2 | 53.9 ± 8.4 | 55.6 ± 7.4 |
| Age-last-exam*** - years | 57.0 ± 8.2 | 60.8 ± 8.5 | 67.3 ± 7.0 |
| time between 1st and last eGFR assessment - median (max) - years | 0.0 (6.1) | 6.8 (40.0) | 11.5 (60.2) |
| Calendar year of exam*** - min to max | 2007 – 2013 | 1970 – 2017 | 1950 – 2017 |
| **eGFR** | | | |
| #eGFR assessments per person – median [IQR] | 1.0 [1.0 - 1.0] | 4.0 [3.0 - 6.0] | 14.0 [11.0 – 19.0] |
| eGFR-first-exam - mL/min/1.73m² | 94.5 ± 13.1 | 96.9 ± 12.1 | 94.3 ± 13.2 |
| eGFR-last-exam - mL/min/1.73m² | 94.3 ± 13.2 | 90.2 ± 12.9 | 81.9 ± 16.3 |
| Chronic Kidney Disease**** – yes (%) | 4,126 (1.6) | 3,871 (3.2) | 15,038 (19.1) |

BMI=Body Mass Index, eGFR=estimated Glomerular Filtration Rate.

* obtained from SC-baseline (data fields 30700, 31, 34, 52, 53, 20116, 21001, 30750, 20003, 2443 and 21001).

** Among the individuals where this variable is available (n=454,361 for smoking status, n=452,721 for BMI).

*** "Exam" is the examination in SC or the examination by the GP, assuming that the date of the eMR-record is the same date as the GP-exam.

**** Chronic Kidney Disease was defined as having at least one assessment of eGFR <60 mL/min/1.73m²

**Supplementary Table 3: Kidney function relevant events**.

We show detailed descriptive statistics across three distinct subsets among individuals in electronic Medical Records (eMR): all data, individuals with a minimum of two eGFR assessments and individuals with at least ten eGFR assessments. In the table header 'n' denotes the number of individuals and 'm' denotes the number of eGFR assessments. We differentiate between prevalent events, reported before the first creatinine assessment and incident event reported after the first recorded creatinine assessments. The results are presented as mean values along with their corresponding standard deviations, if not stated otherwise.

| **Data source** | **All Data (n=199,396)** | **≥2 eGFR assessments (n=195,885)** | **≥10 eGFR assessments (n=69,506)** |
|---|---|---|---|
| **Events before the 1st eGFR assessment** | | | |
| Acute Kidney Injury (%) | 94 (<0.1%) | 94 (<0.1%) | 51 (<0.1%) |
| End-Stage-Kidney-Disease (%) | 49 (<0.1%) | 49 (<0.1%) | 35 (<0.1%) |
| Kidney transplant (%) | 33 (<0.1%) | 33 (<0.1%) | 24 (<0.1%) |
| Dialysis (%) | 30 (<0.1%) | 30 (<0.1%) | 22 (<0.1%) |
| Nephrectomy (%) | 430 (0.2%) | 428 (0.2%) | 236 (0.3%) |
| Pregnancy (%) | 6,003 (3.0%) | 5,848 (3.0%) | 1,208 (1.7%) |
| Diabetes (%) | 15,957 (8.0%) | 15,758 (8.0) | 7,329 (10.5%) |
| **Incident events (i.e. after the 1st eGFR assessment)** | | | |
| Acute Kidney Injury (%) | 563 (0.3%) | 562 (0.3%) | 461 (0.7%) |
| End-Stage-Kidney-Disease (%) | 228 (0.1%) | 228 (0.1%) | 194 (0.3%) |
| Kidney transplant (%) | 180 (<0.1%) | 180 (<0.1%) | 140 (0.2%) |
| Dialysis (%) | 142 (<0.1%) | 142 (<0.1%) | 106 (0.2%) |
| Nephrectomy (%) | 540 (0.3%) | 536 (0.3%) | 352 (0.5%) |
| Pregnancy (%) | 656 (0.3%) | 634 (0.3%) | 148 (0.2%) |
| Diabetes (%) | 15,063 (7.6%) | 14,903 (7.6%) | 7,482 (10.8%) |

**Supplementary Table 4: Overview of variables for time-varying and constant records.**

In our final dataset, we present a set of variables such as the individuals' unique identifier, record-number, record-date and record-type as well as record-specific details that elucidate (**A**) the specific aspects of each time-varying and (**B**) constant records. For each variable, we provide the following information. "Variable Name": The name of the variable. "Type": The data type or format of the variable. "Source": The origin of the variable. "Data Field Description": A brief description of the information encapsulated by the variable.

| Variable Name | Type | Source | Description |
|---|---|---|---|
| **(A) Time-varying records** | | | |
| ID | numeric | SC, eMR | The unique individual identifier from the UK Biobank project number 20272 |
| Record_num | numeric | NA | Unique number of the record |
| Record_date | date | SC, eMR | Date of record |
| Record_type | categorical | NA | Type of record. Can be crea, aki, dialysis, nephrectomy, smoking, bmi, eskd, transplant, diabetes or pregnancy |
| Source | categorical | NA | Source of record. Can be sc or emr |
| Value | numeric | SC, eMR | Value of the record. For example creatinine measure in umol/L if event_type is crea or 1 if event_type is aki |
| **(B) Constant records** | | | |
| ID | numeric | SC | The unique individual identifier from the UK Biobank project number 20272 |
| Date-of-birth | numeric | SC | Date of birth of individual (Data fields 34 and 52) |
| Sex | binary | SC | Sex of individuals (0=female, 1=male, Data field 31) |

.

**Supplementary Figure 1**: **Quality control of serum creatinine measurements from eMRs.**

We extracted all eMR-based serum creatinine measurements recorded in GP-clinical using an approach and a set of read codes provided by Denaxas et al. (*4*). Shown are the number of individuals and measurements (‘n’ and ‘m’ in blue boxes) at various stages of data cleaning. Our cleaning process involved removing zero values of creatinine, eliminating duplicates and same-day measurements, excluding values below the Limit of Detection (LoD) and those exceeding the highest recorded value, and setting values below the Limit of Quantification (LoQ) to the LoQ. Example eGFR values for thresholds of creatinine values were given for a 50-year-old man (*3*).

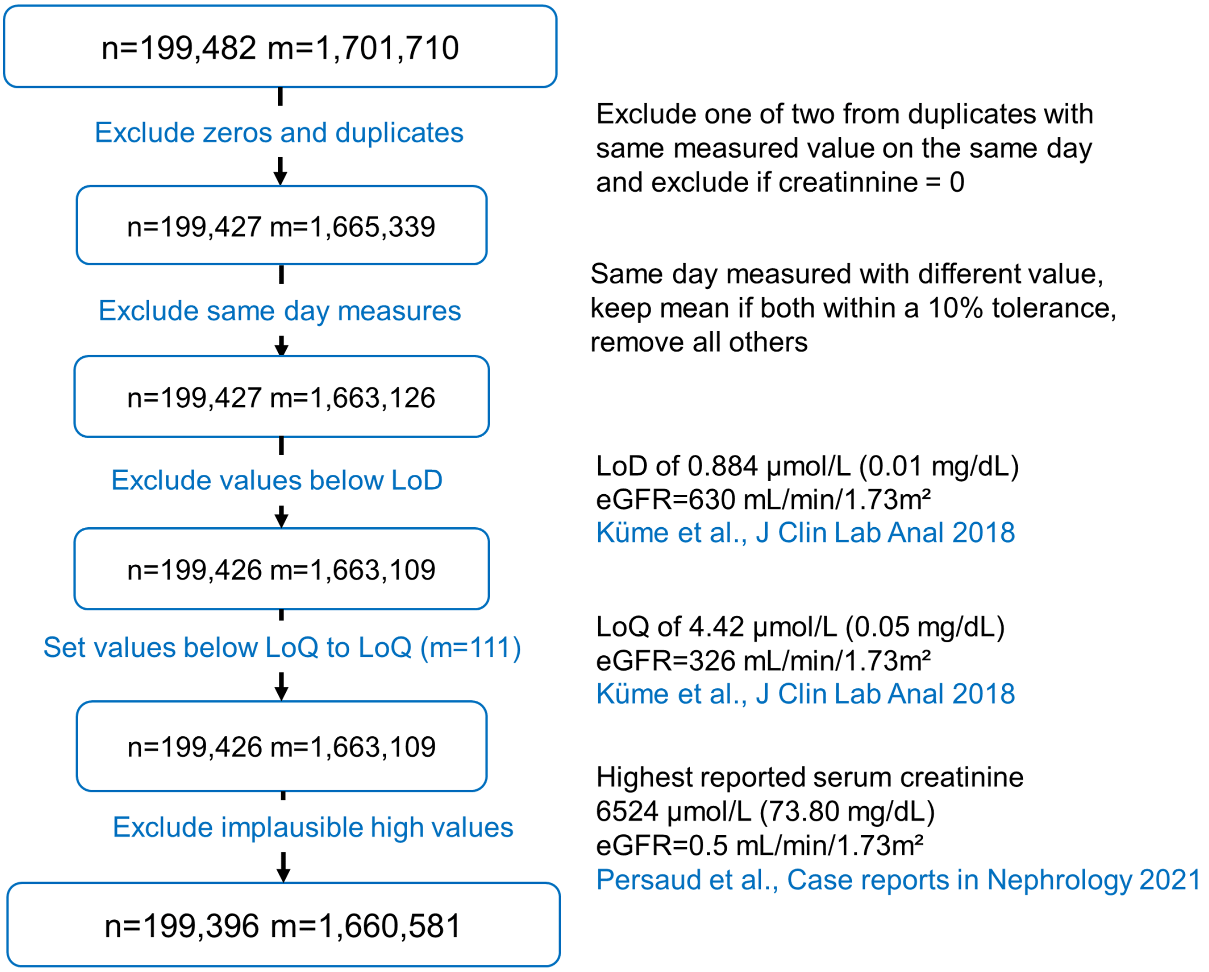

**Supplementary Figure 2: Average between creatinine from Study Center (SC) and electronic Medical Records (eMR)**
By calendar year, we visualize the difference of SC-based and eMR-based creatinine values versus their average in individuals with creatinine from SC and eMR obtained in the same calendar (sample size given above each panel) (*5*). Sex is indicated by color-coding (blue for men, pink for women). Horizontal lines represent mean difference and 95% confidence intervals.

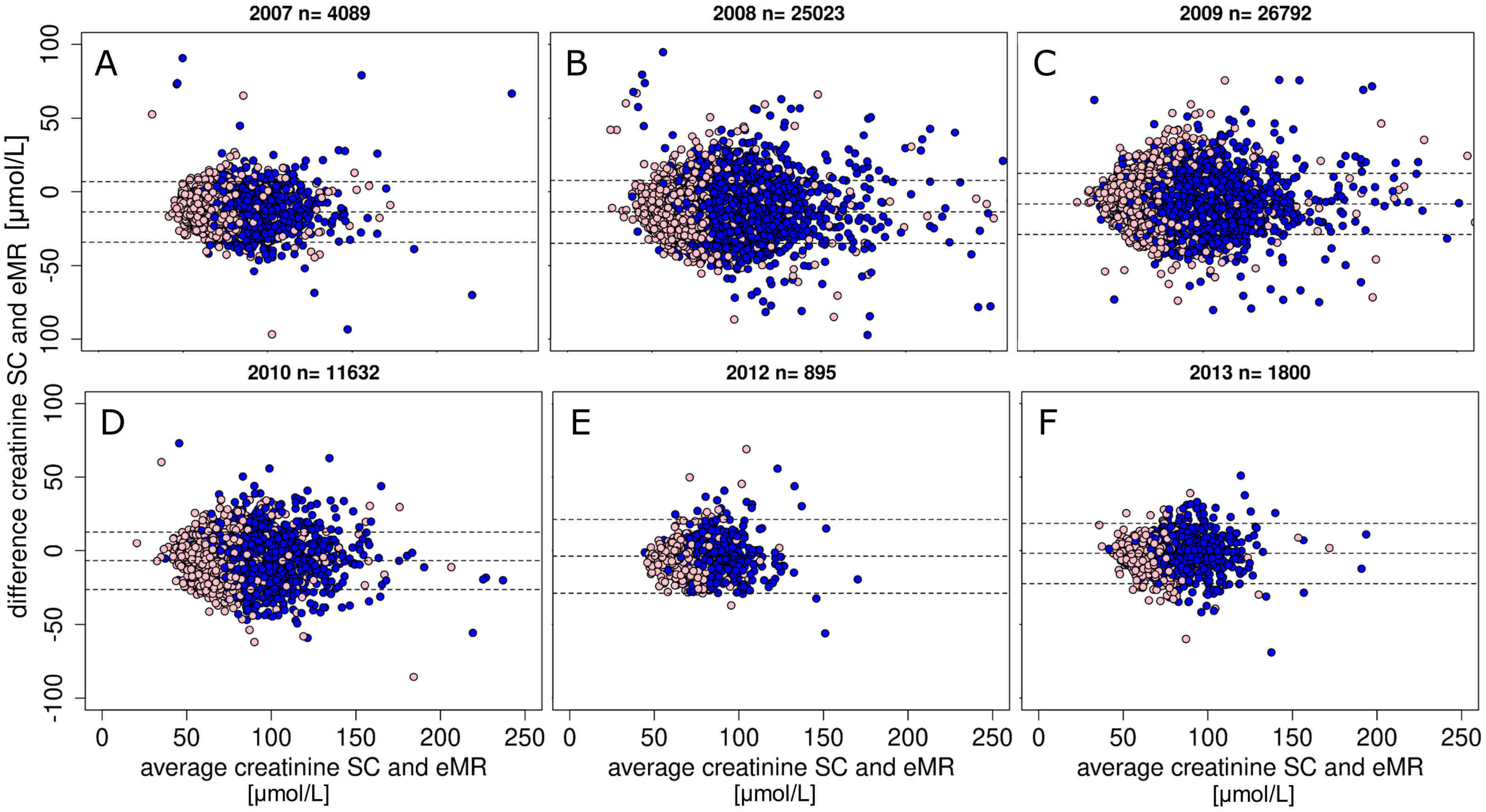

**Supplementary Figure 3: Comparing eGFR values from Study Centre (eGFR$_{SC}$) with eGFR-values from electronic Medical Records (eGFR$_{eMR}$) before correcting the creatinine values from electronic Medical Records**.

We show eGFR$_{SC}$ versus eGFR$_{eMR}$ (quality-controlled, not bias-corrected) among 70,231 individuals with both eGFR-values from the same calendar year (using the eGFR$_{eMR}$ closest to the eGFR$_{SC}$) with gender indicated by color-coding (blue for men and pink for women). We estimated the eGFR via the CKDEPI 2021 formula (*3*). Red dotted lines indicate the identity;

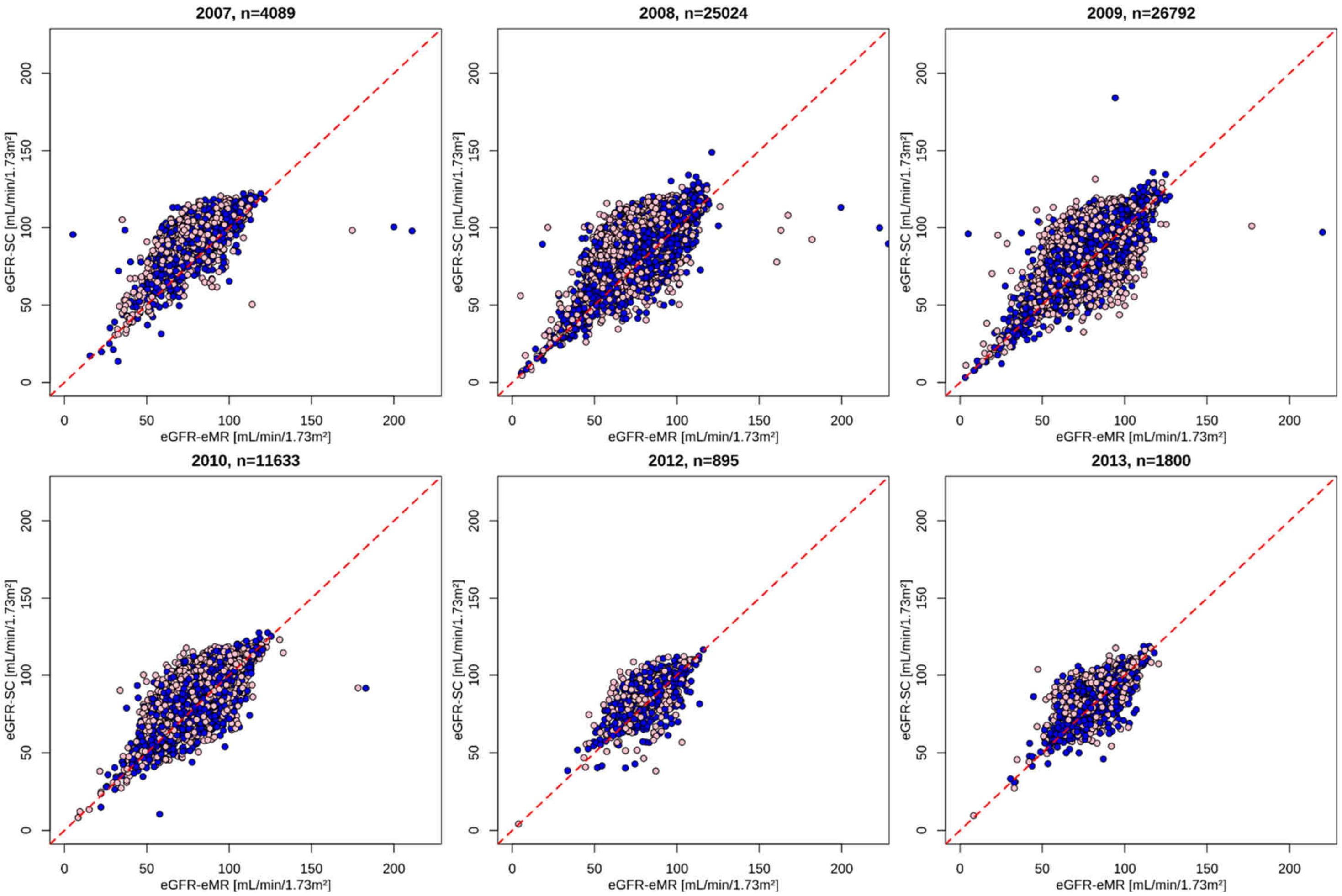

**Supplementary Figure 4: Comparing eGFR values from Study Centre ($\text{eGFR}_{\text{SC}}$) with eGFR-values from electronic Medical Records ($\text{eGFR}_{\text{eMR}}$) after correcting the creatinine values from electronic Medical Records**.

We show $\text{eGFR}_{\text{SC}}$ versus $\text{eGFR}_{\text{eMR}}$ (quality-controlled, bias-corrected) among 70,231 individuals with both eGFR-values from the same calendar year (using the $\text{eGFR}_{\text{eMR}}$ closest to the $\text{eGFR}_{\text{SC}}$) with gender indicated by color-coding (blue for men and pink for women). We estimated the eGFR via the CKDEPI 2021 formula (*3*). Red dotted lines indicate the identity;

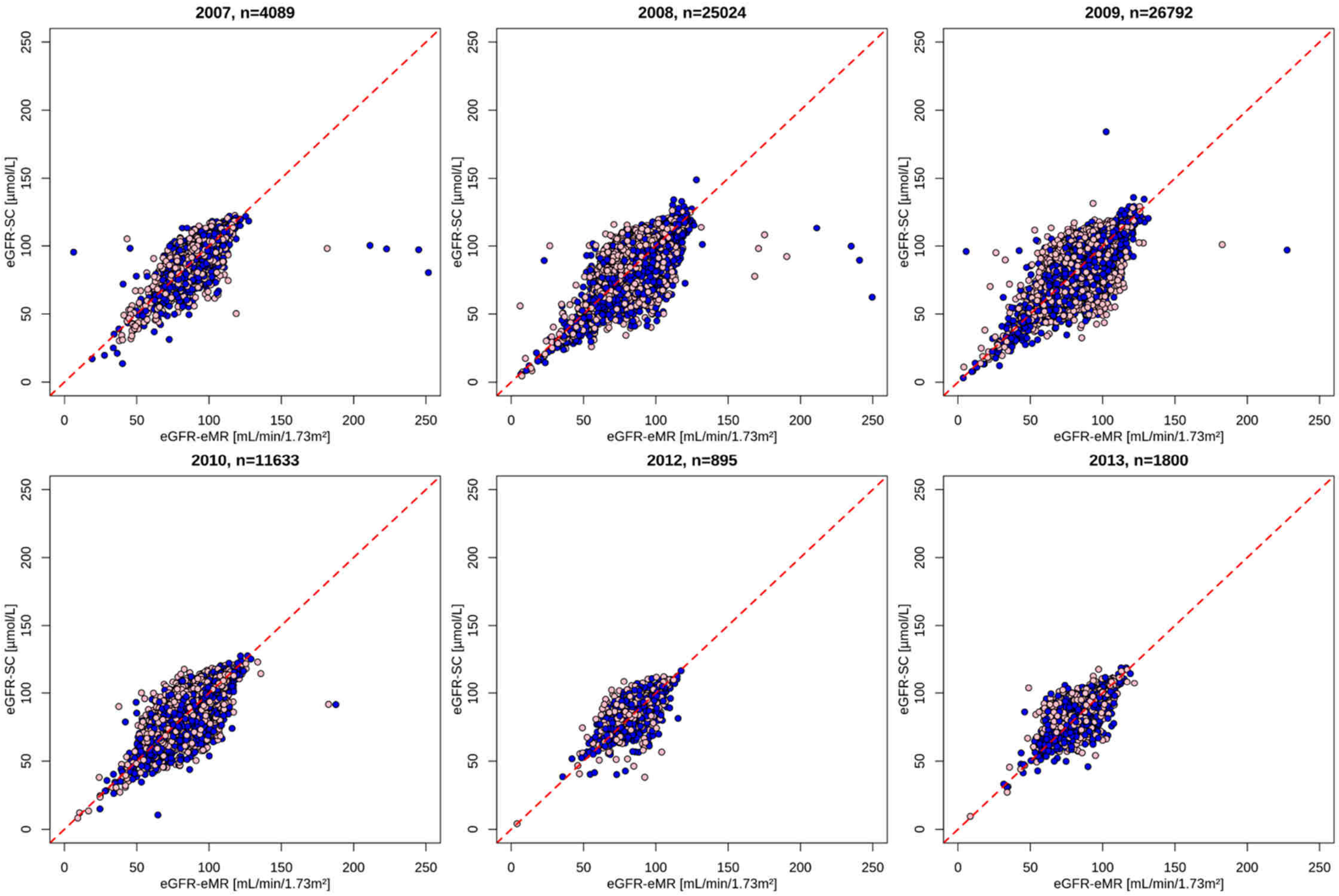

### Supplementary References


1. Küme T, Sağlam B, Ergon C, Sisman AR. Evaluation and comparison of Abbott Jaffe and enzymatic creatinine methods: Could the old method meet the new requirements? J Clin Lab Anal 2018;32.
2. Persaud C, Sandesara U, Hoang V, Tate J, Latack W, Dado D. Highest Recorded Serum Creatinine. Case Reports in Nephrology 2021;2021:1–3.
3. Inker LA, Eneanya ND, Coresh J, Tighiouart H, Wang D, Sang Y, et al. New Creatinine- and Cystatin C-Based Equations to Estimate GFR without Race. N Engl J Med 2021;385:1737–49.
4. Denaxas S, Shah AD, Mateen BA, Kuan V, Quint JK, Fitzpatrick N, et al. A semi-supervised approach for rapidly creating clinical biomarker phenotypes in the UK Biobank using different primary care EHR and clinical terminology systems. JAMIA Open 2020;3:545–56.
5. Bland JM, Altman DG. Statistical methods for assessing agreement between two methods of clinical measurement. Lancet 1986;1:307–10.
